# Beneficial effect of corticosteroids in severe COVID-19 pneumonia: a propensity score matching analysis

**DOI:** 10.1101/2020.05.08.20094755

**Authors:** Tomasz Chroboczek, Marie Lacoste, Chloe Wackenheim, Thibaut Challan-Belval, Benjamin Amar, Thomas Boisson, Jason Hubac, Dominique Leduc, Colleen Masse, Victor Dechaene, Laetitia Touhiri-Maximin, Sandrine Megessier, Camille Lassale

**Author notes:** **Corresponding authors:**, Corresponding author: Tomasz Chroboczek, Mail, Alternate corresponding author: Marie Lacoste, Mail, /.

## Abstract

**Background:** Since December 2019, Severe Acute Respiratory Syndrome Coronavirus 2 (SARS-CoV-2), responsible for Coronavirus Disease 2019 (COVID-19), is spreading worldwide, causing significant morbidity and mortality. No specific treatment has yet clearly shown to improve the disease’s evolution. Validated therapeutic options are urgently needed.

**Methods:** In this retrospective study, we aimed to evaluate determinants of the prognosis of the disease in 70 patients with COVID-19 severe pneumonia (i.e. requiring at least 3 liters of oxygen) hospitalized between 10 March and 9 April, 2020, in the Centre Hospitalier Alpes Léman, France. The main outcome was oro-tracheal intubation and the exposure of interest was corticotherapy. Since this was not a randomized trial, we used propensity score matching to estimate average treatment effect.

**Results:** There was evidence that corticotherapy lowered the risk of intubation with a risk difference of −47.1% (95% confidence interval −71.8% to −22.5%).

**Conclusion:** Corticosteroid, a well-known, easily available, and cheap treatment, could be an important tool in management of severe COVID-19 patients with respiratory failure. Not only could it provide an individual benefit, but also, in the setting of the COVID-19 ongoing pandemic, lower the burden on our vulnerable healthcare systems.

**Key points:** By propensity score matching analysis, the average treatment effect of corticosteroids on 70 patients with severe COVID-19 pneumonia was estimated. Corticosteroid therapy lowered the risk of intubation with a risk difference of −47.1% (95% confidence interval −71.8% to −22.5%).

## Text

Since December 2019, Severe Acute Respiratory Syndrome Coronavirus 2 (SARS-CoV-2), responsible for Coronavirus Disease 2019 (COVID-19), is spreading worldwide, causing significant morbidity and mortality [1-6]. This pandemic has reached at this time more than 187 countries, and the World Health Organization has declared COVID-19 to be a global health concern [7].

The disease has a wide clinical spectrum. Most patients present a mild form of the disease, but some of them are likely to present severe pneumonia, with respiratory failure leading to Acute Respiratory Distress Syndrome, and multiple organ failure leading to death [1-3, 6]. In this process, virally driven aberrant immune-inflammatory response and cytokine storm may play an important role [8-13].

No specific treatment has yet shown clear improvement of the disease’s evolution, due to a lack of high quality studies, and conflicting results. Multiple antiviral treatment options have been evaluated in vitro, but demonstration of their convincing efficacy in vivo needs yet to be done [14-23]. Similarly, several anti-inflammatory drugs, especially corticosteroids and tocilizumab, are candidates to treat patients with COVID-19 pneumonia [9, 12, 13, 24-28]. This type of treatment may be useful in case of administration after viral clearance, during the inflammatory phase of the disease, in order to dissipate the “cytokine storm” that can occur following the viral trigger in the setting of aberrant immune-inflammatory response [9, 24-27]. Overall, studies to validate new therapeutic options are urgently needed.

In this study, we sought to evaluate which parameters can be associated with the prognosis of the disease in 70 patients with COVID-19. In particular, we wanted to assess the effect of corticosteroids in the case of severe pneumonia.

## METHODS

We investigated retrospectively all cases of COVID-19 in adult patients (i.e. older than 18 years), hospitalized between 10 March and 9 April, 2020, in the Centre Hospitalier Alpes Léman, in Contamine sur Arve, France. Each case was defined as an association of clinical symptoms attributed to SARS-CoV-2 infection with the isolation of specific RNA by RT-PCR on naso-pharyngeal swab. Of all the COVID-19 patients, we identified a subset of patients with respiratory failure due to pneumonia (diagnosed by chest imaging), and requiring more than 3 liters of oxygen to obtain oxygen saturation higher than 92%, at any time of hospitalization. Since we wanted to study factors associated with oro-tracheal intubation, we excluded from the study patients for whom oro-tracheal intubation was advised against, after collegial discussion.

The following data were collected: demographic information, medical history, clinical characteristics, laboratory data, radiologic report data, comorbidities, therapeutic interventions during the hospitalization, and clinical outcomes. The patient’s demographic information (age and gender), clinical characteristics (fever, cough, dyspnea and oxygen dose requirement at day 1 of hospitalization, dysosmia, dysgeusia), and duration from symptom’s onset (i.e. time point when the symptoms were first noticed by patients or their relatives) to admission were collected from the electronic medical system. The radiologic report data (i.e. damages diagnosed with chest computed tomography (CT) Scan) were obtained from the communication system. Laboratory data (blood cell count, C-reactive protein (CRP), organ function markers) were collected from the laboratory information system. Comorbidities (cardio-vascular disease, including hypertension, diabetes, chronic renal diseases, overweight with calculation of Body Mass Index) were extracted from medical history, and classified according to the Charlson Comorbidity Index. In-hospital medications and interventions were also collected from the electronic medical system. If the patient got intubated and ventilated (outcome of interest), we took into account medication and interventions that started at least 24 hour prior to intubation. Each participant was given a study number using an electronic coding system before data extraction in order to preserve the patient’s anonymity.

The study received approval from the local Ethics Committee of the Centre Hospitalier Alpes Léman. In accordance with French law, every patient was informed of the exploitation of their medical information in this non-interventional and retrospective study, and their right to formulate opposition to its use. The study was declared to the National Comity for Informatics and Liberties (CNIL), as required by law.

Description of patients’ characteristics was expressed as means and standard deviation (SD) for continuous variables, and number (percentage) for categorical variables. To establish the factors associated with oro-tracheal intubation, patient demographics, clinical presentation, diagnostic test results, treatments, and disease progression (before intubation for the ventilated patients) were compared between ventilated and non-ventilated patients. Univariate comparisons were performed using Fisher’s exact test, or chi-square test for categorical data, and independent Student’s t-tests for continuous data, as appropriate. Following univariate analysis, the independent contribution of patient’s characteristics to the risk of oro-tracheal intubation was analyzed. First, we did a multivariate logistic regression to model the odd ratios of intubation with all variables of interest in the model. Considering comorbidities, we chose to evaluate the impact of Charlson Comorbidity Index, high blood pressure, and BMI; other comorbidities were considered redundant and therefore excluded from the model. Impact of severity was assessed by including the oxygen dose at entrance to the hospital in the model, and the highest CRP level was used to evaluate the impact of systemic inflammation. Medications were also included in the model. Odds ratios (OR) were estimated with 95% confidence intervals (95% CI). Statistical significance was defined as a *p* value less than 0.05.

To specifically assess the effect of corticosteroid treatment on risk of oro-tracheal intubation and to overcome indication bias, we used a propensity score matching method to estimate average treatment effect (ATE). We created a propensity score of exposure to corticosteroids using a logistic regression model including the following covariates: age, sex, Charlson index (categorical >1), BMI>25 kg/m^2^, time from initiation of symptoms to hospitalization >7 days, CRP>150 mg/L, hypertension, oxygen>3L, treatment with hydroxychloroquine, treatment with azithromycin. These variables include potential confounders (related to treatment and outcome) and prognosis of the outcome [29]. The ATE evaluates the average effect of a treatment in a population where all individuals have the same probability of receiving the treatment. It compares the effect on the outcome (i.e. here oro-tracheal intubation) of the treated versus the untreated patients. Statistical analyses were performed with the Stata version 15.1 program (StataCorp, College Station, TX).

## RESULTS

### Patients’ characteristics

Between March 10 and April 9, 2020, 267 adult patients were hospitalized for COVID-19 in the Centre Hospitalier Alpes Léman, in Contamine sur Arve, France, both in Medicine departments, and Intensive Care Units (ICU). Among these, 34 patients were excluded from our study due to a collegial decision of health care limitation. Upon the 233 remaining patients, 70 (30%) required more than 3 l/mn of oxygen during their hospitalization in order to reach saturation above 93%. These 70 cases were investigated, and their characteristics are presented in **Table 1**.

**Table 1.**
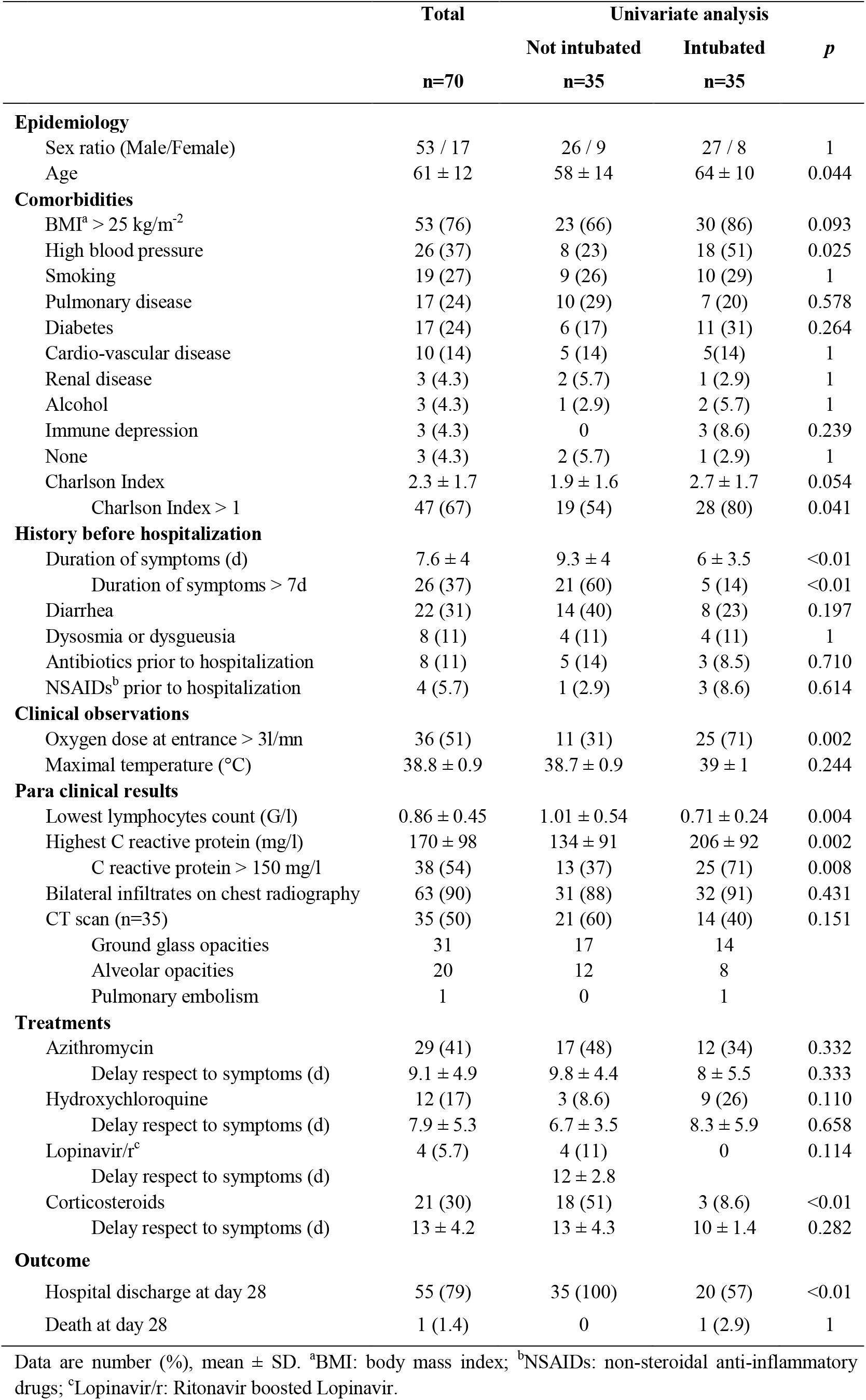
Patients’ characteristics according to their evolution.

Patients were mainly men (53, versus only 17 women), with a mean age of 61 ± 12 years. Only 3 (4.3%) of them had no comorbidity. Being overweight (i.e. BMI > 25) was the main comorbidity (76%). High blood pressure, smoking history, pulmonary comorbidity, and diabetes were also frequent, with 26 (37%), 19 (27%), 17 (24%) and 17 (24%) cases, respectively. The mean duration of symptoms before hospitalization was 7.6 ± 4 days. Extra-respiratory symptoms could be present, with frequent diarrhea (22 cases, 31%), as well as dysosmia or dygueusia (8 cases, 11 %). The mean maximal temperature during hospitalization was 38.8 ± 0.9 °C. Fifty-one (73%) patients had a lowest lymphocyte count above 1 g/L, with a mean lowest lymphocytes count of 0.86 ± 0.45 G/L. Mean highest C Reactive Protein (CRP) was 170 ± 98 mg/L. Only 35 (50%) patients underwent a thoracic CT scan. Notably, ground glass opacities were not constant (31 of 35 cases, 89%).

Three different assumed antiviral treatments were prescribed at our institution: Azithromycin, Hydroxychloroquine, and Ritonavir-boosted Lopinavir (Lopinavir/r) were used in 29 (41%), 12 (17%) and 4 (5.7%) cases, respectively. Corticosteroids were given in 21 (30%) cases. Delays of instauration of these treatments after symptom’s onset were 9.1 ± 4.9 days, 7.9 ± 5.3 days, 12 ± 2.8 days, and 13 ± 4.2 days, respectively. At day 28 after admission to hospital, 55 (79%) patients were discharged from the hospital, and 1 (1.4%) was dead.

### Patients’ characteristics according to their evolution

Thirty-five (50%) patients required oro-tracheal intubation and mechanical ventilation during hospitalization, due to respiratory failure in every case. Age, BMI ≥ 25 kg/m^-2^, high blood pressure, Charlson index > 1, duration of symptoms, oxygen dose > 3 L/min, lowest lymphocytes count and highest CRP, and administration of corticosteroids, were statistically significantly different between patients who underwent mechanical ventilation and those who did not (**Table 1**).

The multivariable analysis showed that an oxygen dose > 3 L/min at admission, a highest CRP >150 mg/l, and administration of hydroxychloroquine were independently associated with the risk of intubation, whereas corticosteroid therapy had a protective effect (p<0.05). Results are presented in **Table 2**.

**Table 2:**
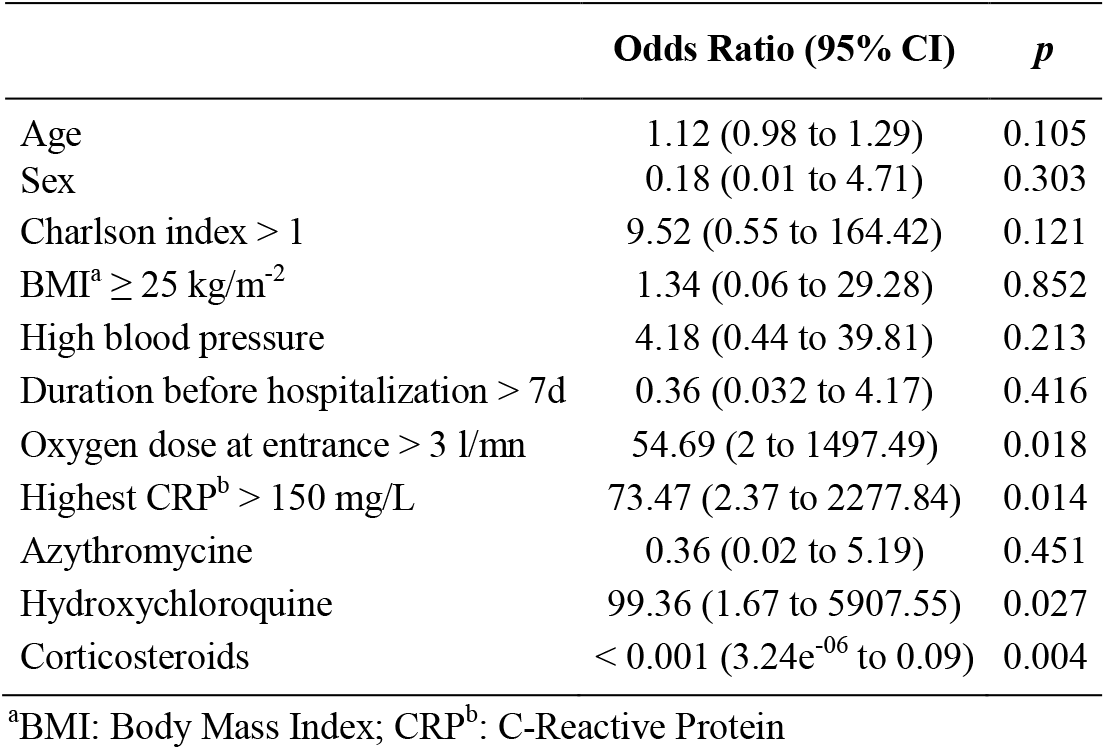
Multivariable odds ratios of intubation associated with patients characteristics, n=70.

### Corticosteroids effect

We created a propensity score model to adjust and balance the groups (i.e. corticosteroids treated and untreated patients), using matching method, in order to predict the exposure to corticosteroids and to estimate the ATE. We found that corticosteroid therapy affected the risk of intubation with a risk difference of −47.1% (95% CI −71.8 to −22.5).

## DISCUSSION

In this study, we showed that corticosteroids prescription is likely to prevent the worsening of COVID-19 severe pneumonia, as its administration was associated with a lower risk of oro-tracheal intubation and ventilation in the ICU. This result is of primary importance, at the time of an ongoing pandemic, with a healthcare system that is overwhelmed in many regions of the world.

For instance, in our hospital, between March 10 and April 9, 2020, among the 70 patients that required more than 3 L/mn of oxygen, 35 were intubated and ventilated. In our sample, 65% of patients who did not receive corticosteroid therapy were intubated. Systematic prescription of corticosteroids to all these patients may have resulted, based on our results, in 17.9% (= 65% - 47.1%) of intubations, i.e. 12 intubations overall. In our hospital, the ICU is usually composed of 15 beds only; at the peak of the first wave of the pandemic, it required deep reorganization, and extension of its capacity to 35 beds, due to the strong increase in the number of COVID-19 ventilated patients. With more frequent prescription of corticosteroids, the pressure upon our hospital would likely have been much lower. At a higher level, the same logic can be applied to French healthcare system as a whole: at the beginning of April 2020, at the peak of the first wave, more than 7000 people were hospitalized in French ICUs [30], with important regional disparities in disease incidence, whereas the whole system’s capacity consists of 7000 beds. This has resulted in important disorganization of the system, and a likely increase in morbi-mortality and costs [31]. Between 1 March and 13 April, 2020, 3 French departments (Haut-Rhin, Seine-Saint-Denis, and Haut-De-Seine) deplored more than 2 times more deaths than in either 2019 or 2018 [31]. Considering countries with less prepared healthcare systems, the benefits of corticotherapy could be even more significant.

At this time, comparative studies describing anti-inflammatory treatment of COVID-19 are scarce. Zha et al. restrospectively described 31 patients with mild COVID-19 (only 4 presented dyspnea), of which 11 received corticosteroids treatment. They found, in multivariate analysis, no difference in terms of viral clearance, length of hospital stay, or duration of symptoms [26]. Wang et al. retrospectively reviewed 46 hospitalized patients with COVID-19 pneumonia, of which 26 received corticosteroid treatment. There was no information about the severity of respiratory failure, but average saturation at entrance of 91%, with interquartile range going from 86 to 92%. Comparison in univariate analysis between patients with and without corticosteroids showed a shorter duration of fever, and a faster improvement in saturation in the corticosteroid group [27]. Roumier et al. retrospectively compared 30 patients with severe COVID-19 pneumonia, i.e. requiring more than 6 liters of oxygen, treated with tocilizumab, with 30 patients matched for age, gender, and disease severity. Treatment with tocillizumab significantly reduced the risk of subsequent ICU admission [25]. Overall, based on these limited results, one could put forth the assumption that the higher the oxygen necessity and the higher the inflammation, the more effective anti-inflammatory drugs could be.

In our study, mean duration of symptoms before hospitalization was 7.6 ± 4 days; the introduction of the different drugs therefore occurred mainly after the first week of disease evolution, both for antiviral drugs, and for corticosteroids. Our team’s consensus about the timing of corticosteroid prescription was to always wait for at least 7 days after the onset of symptoms, supposedly in order to avoid a deleterious effect on viral clearance and to limit the inflammatory phase in the same time. We could not find any information about the timing of the prescription with respect to symptoms onset in the 3 comparative studies mentioned above [25-27].

Not only did we find no beneficial effect of Azithromycin or Hydroxychloroquine administration, but Hydroxychloroquine was significantly associated with higher odds of intubation. This association might be explained by a real deleterious effect of this treatment, that has already been shown elsewhere [14, 18]. The question of the effect of this treatment according to the delay from the onset of symptoms still needs to be clarified. However, it is very important to note that, in our hospital, Hydroxychloroquine was prescribed more often to patients hospitalized in ICUs, i.e. with greater disease severity. Therefore, the observed association between hydroxychloroquine and higher odds of intubation may be the result of prescription bias. Nevertheless, the inclusion of an indicator of severity in the multivariable analysis (requiring > 3L/mn of oxygen at study entry) should have minimized this prescription bias, although unmeasured confounding cannot be ruled out. Caution in interpreting this OR is advised, as the study of the effect of hydroxychloroquine was not the aim of the present study. However, its inclusion in the propensity score model was mandatory, since it is a variable related to the outcome [29].

Our study presents numerous limitations, notably those that are inherent to its retrospective nature. Nonetheless, we tried to clarify the role of corticosteroids on the disease’s evolution by building a propensity score, and thus limit confounding due to prescription bias. Also, we chose to consider and investigate all patients hospitalized for COVID-19 in our institution, to limit the selection bias; however, this could have induced prescription bias, since not all the hospital units had the same prescription practice. We did not notice any serious adverse effect of corticosteroid therapy in our study, but it is worth noting that long-term effects cannot be investigated at this time. This should be the object of further studies.

In conclusion, we believe that corticosteroid, a well-known, easily available, and cheap treatment, could be an important tool for the management of severe COVID-19 patients with respiratory failure. Not only could it provide an individual benefit, but also, in the context of the current COVID-19 pandemic, lower the burden on our vulnerable healthcare systems.

## Data Availability

The data that support the findings of this study are available from the corresponding author, T.C., upon reasonable request.

## Acknowledgments

Authors are deeply indebted to Juliusz Chroboczek for his help with proof reading.

## Contribution

All authors have seen and approved the manuscript, and contributed significantly to the work.

## References

1. Du Y, Tu L, Zhu P, et al. Clinical Features of 85 Fatal Cases of COVID-19 from Wuhan: A Retrospective Observational Study. Am J Respir Crit Care Med 2020.

2. Chen N, Zhou M, Dong X, et al. Epidemiological and clinical characteristics of 99 cases of 2019 novel coronavirus pneumonia in Wuhan, China: a descriptive study. Lancet 2020; 395(10223): 507–13.

3. Wang Z, Yang B, Li Q, Wen L, Zhang R. Clinical Features of 69 Cases with Coronavirus Disease 2019 in Wuhan, China. Clin Infect Dis 2020.

4. Zhu N, Zhang D, Wang W, et al. A Novel Coronavirus from Patients with Pneumonia in China, 2019. N Engl J Med 2020; 382(8): 727–33.

5. Zhou P, Yang XL, Wang XG, et al. A pneumonia outbreak associated with a new coronavirus of probable bat origin. Nature 2020; 579(7798): 270–3.

6. Wang D, Hu B, Hu C, et al. Clinical Characteristics of 138 Hospitalized Patients With 2019 Novel Coronavirus-Infected Pneumonia in Wuhan, China. Jama 2020.

7. Johns Hopkins University and Medicine. COVID-19 Dashboard by the Center for Systems Science and Engineering (CSSE) at Johns Hopkins University (JHU). Available at: https://coronavirus.jhu.edu/map.html.

8. Liu F, Li L, Xu M, et al. Prognostic value of interleukin-6, C-reactive protein, and procalcitonin in patients with COVID-19. J Clin Virol 2020; 127: 104370.

9. Zhang S, Li L, Shen A, Chen Y, Qi Z. Rational Use of Tocilizumab in the Treatment of Novel Coronavirus Pneumonia. Clin Drug Investig 2020.

10. Chen X, Zhao B, Qu Y, et al. Detectable serum SARS-CoV-2 viral load (RNAaemia) is closely correlated with drastically elevated interleukin 6 (IL-6) level in critically ill COVID-19 patients. Clin Infect Dis 2020.

11. Mehta P, McAuley DF, Brown M, Sanchez E, Tattersall RS, Manson JJ. COVID-19: consider cytokine storm syndromes and immunosuppression. Lancet 2020; 395(10229): 1033–4.

12. Xu X, Han M, Li T, et al. Effective treatment of severe COVID-19 patients with tocilizumab. Proceedings of the National Academy of Sciences 2020: 202005615.

13. Sciascia S, Apra F, Baffa A, et al. Pilot prospective open, single-arm multicentre study on off-label use of tocilizumab in patients with severe COVID-19. Clin Exp Rheumatol 2020.

14. Barbosa J, Kaitis D, Freedman R, Le K, Lin X. Clinical Outcomes of Hydroxychloroquine in Hospitalized Patients with COVID-19: A QuasiRandomized Comparative Study. Submitted to the New England Journal of Medicine. Available at: https://www.dropbox.com/s/urzapkyij542qx5/NEJM_Clinical%20Outcomes%20of%20Hydrox140ychlorquine%20in%20Patients%20with%20COVID19.pdf.pdf.pdf.pdf.pdf.pdf.pdf.pdf?dl=0

15. Chen Z, Hu J, Zhang Z, et al. Efficacy of hydroxychloroquine in patients with COVID-19: results of a randomized clinical trial. medRxiv 2020: 2020.03.22.20040758.

16. Gautret P, Lagier JC, Parola P, et al. Hydroxychloroquine and azithromycin as a treatment of COVID-19: results of an open-label non-randomized clinical trial. Int J Antimicrob Agents 2020: 105949.

17. Gautret P, Lagier JC, Parola P, et al. Clinical and microbiological effect of a combination of hydroxychloroquine and azithromycin in 80 COVID-19 patients with at least a six-day follow up: A pilot observational study. Travel Med Infect Dis 2020: 101663.

18. Magagnoli J, Narendran S, Pereira F, et al. Outcomes of hydroxychloroquine usage in United States veterans hospitalized with Covid-19. medRxiv 2020: 2020.04.16.20065920.

19. Yu B, Wang DW, Li C. Hydroxychloroquine application is associated with a decreased mortality in critically ill patients with COVID-19. medRxiv 2020: 2020.04.27.20073379.

20. Wang Y, Zhang D, Du G, et al. Remdesivir in adults with severe COVID-19: a randomised, double-blind, placebo-controlled, multicentre trial. The Lancet 2020.

21. Borba MGS, Val FFA, Sampaio VS, et al. Effect of High vs Low Doses of Chloroquine Diphosphate as Adjunctive Therapy for Patients Hospitalized With Severe Acute Respiratory Syndrome Coronavirus 2 (SARS-CoV-2) Infection: A Randomized Clinical Trial. JAMA Netw Open 2020; 3(4): e208857.

22. Cai Q, Yang M, Liu D, et al. Experimental Treatment with Favipiravir for COVID-19: An Open-Label Control Study. Engineering (Beijing) 2020.

23. Cao B, Wang Y, Wen D, et al. A Trial of Lopinavir-Ritonavir in Adults Hospitalized with Severe Covid-19. N Engl J Med 2020.

24. Radbel J, Narayanan N, Bhatt PJ. Use of tocilizumab for COVID-19 infection-induced cytokine release syndrome: A cautionary case report. Chest 2020.

25. Roumier M, Paule R, Groh M, Vallee A, Ackermann F. Interleukin-6 blockade for severe COVID-19. medRxiv 2020: 2020.04.20.20061861.

26. Zha L, Li S, Pan L, et al. Corticosteroid treatment of patients with coronavirus disease 2019 (COVID-19). Med J Aust 2020.

27. Wang Y, Jiang W, He Q, et al. Early, low-dose and short-term application of corticosteroid treatment in patients with severe COVID-19 pneumonia: single-center experience from Wuhan, China. medRxiv 2020: 2020.03.06.20032342.

28. Gritti G, Raimondi F, Ripamonti D, et al. Use of siltuximab in patients with COVID-19 pneumonia requiring ventilatory support. medRxiv 2020: 2020.04.01.20048561.

29. Brookhart MA, Schneeweiss S, Rothman KJ, Glynn RJ, Avorn J, Sturmer T. Variable selection for propensity score models. Am J Epidemiol 2006; 163(12): 1149–56.

30. Santé Publique France. Géo Données en Santé Publique. Available at: https://geodes.santepubliquefrance.fr/#c=indicator&f=0&i=covid_hospit_clage10.rea&s=2020-05-02&t=a01&view=map1.

31. Institut National de la Statistique et des Etudes Economiques (INSEE). Nombre de décès quotidiens par département. Available at: https://www.insee.fr/fr/information/4470857.

